# Severe acute respiratory syndrome coronavirus 2 breakthrough infections in healthcare workers vaccinees with BNT162b2 (Pfizer-BioNtech) in Bogotá, Colombia

**DOI:** 10.1101/2022.05.31.22274501

**Authors:** Pilar Tavera-Rodríguez, Juliana Barbosa-Ramírez, Andrea Bermúdez-Forero, Diego Prada-Cardozo, Jhonnatan Reales-González, Dioselina Peláez-Carvajal, Diana Malo-Sanchez, Maria-Ximena Meneses-Gil, Marcela Mercado-Reyes

## Abstract

The healthcare workers are considered as a high-risk group for infection with SARS-CoV-2, so they were included in the first stage of the National Plan for Vaccination against COVID-19 in Colombia.

An ongoing prospective cohort study to evaluate immune response to vaccination included 490 workers from health institutions in Bogotá, Colombia, vaccinated between March and June 2021 with BNT162b2 (Pfizer-BioNtech). Multiple samples were collected during a follow-up period of 6 months after immunization. We report cases of asymptomatic and symptomatic SARS-CoV-2 infections detected in this cohort. For each participant demographic data, vaccination dates, results for SARS-CoV-2 RT-PCR, and detection of antibody (IgG) tests during the follow-up period were collected.

SARS-CoV-2 infection was detected in 38 (7.7 %) volunteers. Of these, 81.6% had a positive RT-PCR for SARS-CoV-2, and 18.4% were confirmed by detection of IgG anti-SARS-CoV-2 nucleoprotein; 76.3% of infections occurred after 7 days of second dose. A total of 57.9% of the cases were asymptomatic. No hospitalizations or deaths were registered. When infection occurred, 81.6% of infected participants had presence of IgG anti-S antibodies. In 12 samples in which genomic characterization was achieved, 83.4% corresponded to the variant Mu, 8.3% Gamma, and 8.3% Delta.

All findings agree with other reports in different studies that show the benefit of COVID-19 vaccines, protecting specially against severe disease but not against infection or re-infection.

## Background

In Colombia the National Plan for Vaccination against coronavirus disease 2019 (COVID-19) started on February 17 2021, with healthcare workers because of the high risk of exposure, using the BNT162b2 COVID-19 vaccine manufactured by Pfizer and BioNTech; healthcare workers at every level and service were included [1]. There are multiple reports about BNT162b2 vaccine efficacy in preventing symptomatic severe acute respiratory syndrome coronavirus 2 (SARS-CoV-2) infections, complications due to infection, and deaths with values above 85% and 90% for symptomatic infection and deaths, respectively [2 - 5]. However, vaccinated individuals are still at risk of infection.

## Objectives

To report SARS-CoV-2 cases with asymptomatic and symptomatic infections in a cohort of healthcare workers detected during 6–9 months of follow-up after vaccination with BNT162b2.

## Study design

An ongoing prospective cohort study to evaluate the humoral immune response to vaccination included 490 workers from health institutions in Bogotá, Colombia, vaccinated between March and June 2021 with BNT162b2 (Pfizer-BioNTech). Five follow-ups were carried out in 6 months (baseline 0–3 days before vaccination, day 21, 60, 90, and 210 after the first dose) and included the collection of blood samples and a nasopharyngeal swab from participants. Additional follow-up was performed on volunteers who reported symptoms compatible with COVID-19 or close contact with confirmed cases until 9 months after vaccination.

This study presents information on cases of SARS-CoV-2 infection detected in the cohort. In addition, for each participant, demographic data, vaccination dates, results for SARS-CoV-2 reverse transcription-polymerase chain reaction (RT-PCR), and detection of antibody (immunoglobulin G [IgG]) tests were collected. The study was approved by the National Health Institute Ethics Committee (CEMIN-4-2021), and all participants signed the informed consent. Anti-S1 IgG was quantified using a SARS-CoV-2 IgG sCOVG chemiluminescence immunoassay (ADVIA Centaur XPT platform, Siemens). In addition, anti-SARS-CoV-2 nucleocapsid IgG presence were determined using the ID Screen®SARS-CoV-2-N indirect enzyme-linked immunosorbent assay (ID Vet).

SARS-CoV-2 was detected in nasopharyngeal swabs using real-time RT-PCR to detect the viral E gene [6]. Positive samples by RT-PCR with a cycle threshold (CT) <25 were included in sequencing [7], assembly of raw data [8], and lineage assignment [9].

## Results

SARS-CoV-2 infection was detected in 38 (7.7 %) participants during the follow-up period, with a median age of 40 years (range 26–62); 65.8% (25/38) of them were women. Administrative and laboratory staff had the same frequency (32%), followed by field epidemiologists (29%) and general service staff (8%).

The 78.9% (30/38) of positive cases occurred between May and July 2021; therefore, cases of COVID-19 in healthcare workers described here occurred during the third peak of the disease in Colombia (Figure 1).

**Figure 1.**
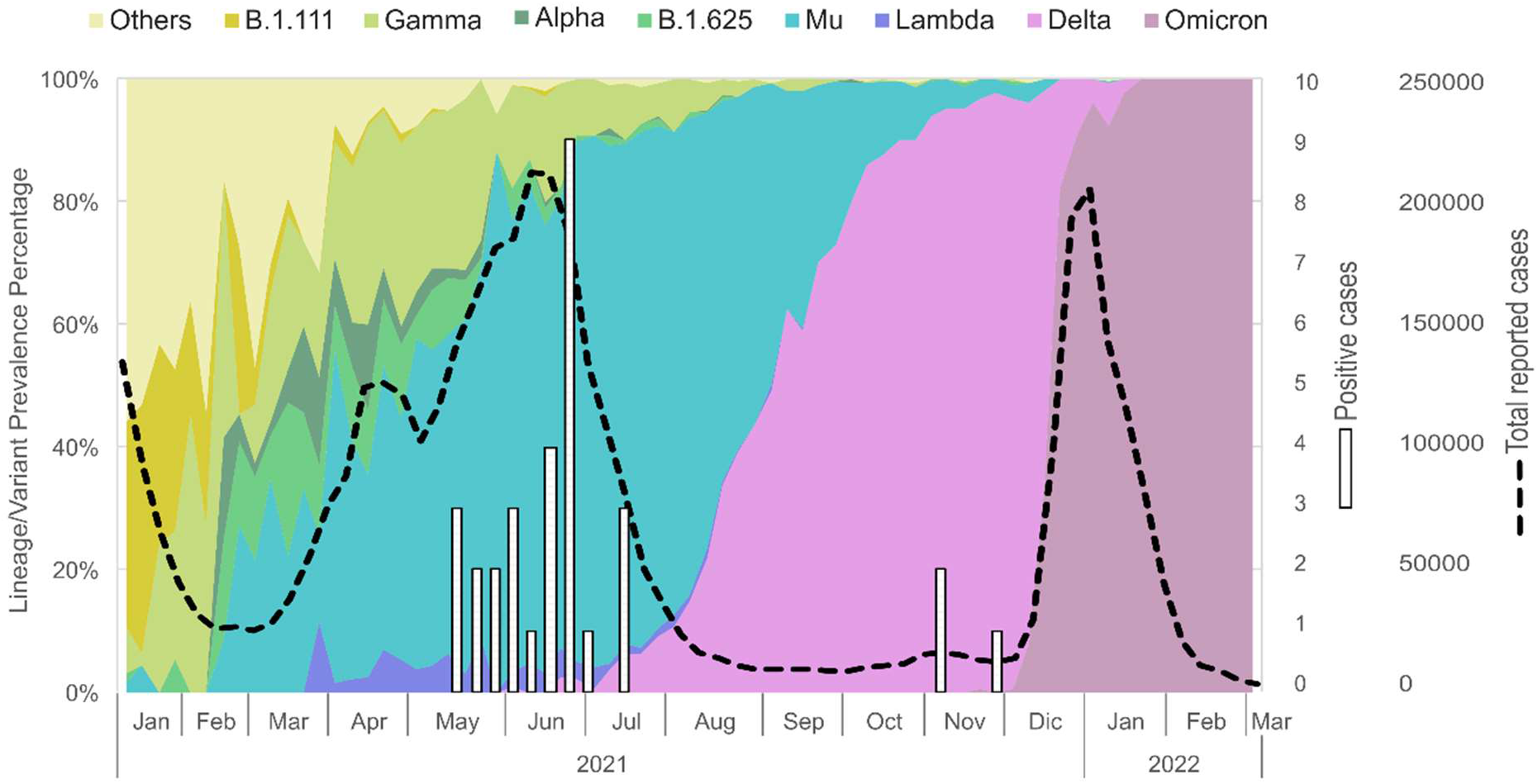
Control graph incidence of confirmed SARS-CoV-2 cases, 2021*. * Cases shown corresponds to those confirmed by RT-PCR because cases confirmed by serology do not have a specific date of infection or symptoms onset.

A total of 22 out of 38 were asymptomatic. Between the remaining 16 cases, the most frequent symptoms were adynamia (62.5%), headache (43.8%), and fever (37.5%) (Table 1). No hospitalizations or deaths were register.

**Table 1.** Information for cases of SARS-CoV-2 infection confirmed by RT-PCR.

| Case | Sex / Age range | Occupation / activity | Date of positive result (Epi week - year) | Days after 1st dose | Symptoms | Variant / Lineage | Anti-S IgG (BAU/ml)<sup>§</sup> |
| --- | --- | --- | --- | --- | --- | --- | --- |
| 1 | F/<br>25-34 | Field epidemiology | 21 - 2021 | 78 | Cough, runny nose, fever, headache, adynamia, ageusia | Mu (B.1.621) | >2180 |
| 2 | F/<br>25-34 | Field epidemiology | 20 - 2021 | 74 | Asymptomatic | Gamma (P.1) | >2180 |
| 3 | M/<br>35-44 | Laboratory | 19 - 2021 | 0 | Asymptomatic | Mu (B.1.621) | No Reactive |
| 4 | M/<br>45-54 | General services | 19 - 2021 | 0 | Asymptomatic | Mu (B.1.621) | 288.9 |
| 5 | F/<br>45-54 | General services | 19 - 2021 | 0 | Fever, adynamia, myalgia | NS | No Reactive |
| 6 | F/<br>35-44 | Laboratory | 22 - 2021 | 34 | Asymptomatic | NS | 670.6 |
| 7 | F/<br>35-44 | Administrative | 22 - 2021 | 20 | Asymptomatic | NS | 93.1 |
| 8 | F/<br>25-34 | Administrative | 22 - 2021 | 20 | Headache, adynamia, anosmia, ageusia | Mu (B.1.621) | 56.7 |
| 9 | F/<br>25-34 | Laboratory | 23 - 2021 | 95 | Asymptomatic | NS | 570.7 |
| 10 | M/<br>45-54 | Administrative | 20 - 2021 | 8 | Asymptomatic | NS | No Reactive |
| 11 | M/<br>35-44 | Laboratory | 21 - 2021 | 14 | Headache, adynamia, fever, arthralgia, eye pain, cough, fatigue | NS | No Reactive |
| 12 | M/<br>45-54 | Laboratory | 24 - 2021 | 32 | Asymptomatic | Mu (B.1.621) | No Reactive |
| 13 | F/<br>45-54 | Administrative | 24 - 2021 | 10 | Headache, adynamia, anosmia | NS | No Reactive |
| 14 | M/<br>55-64 | Administrative | 24 - 2021 | 35 | Fever, sore throat | Mu (B.1.621) | >2180 |
| 15 | F/<br>35-44 | Laboratory | 24 - 2021 | 103 | Asymptomatic | NS | 2775,1 |
| 16 | F/<br>35-44 | Administrative | 25 - 2021 | 72 | Asymptomatic | Mu (B.1.621) | >2180 |
| 17 | F/<br>35-44 | Laboratory | 25 - 2021 | 40 | Asymptomatic | Mu (B.1.621) | 222.1 |
| 18 | M/<br>45-54 | Administrative | 25 - 2021 | 41 | Adynamia, runny nose, coryza, myalgia | NS | 87.2 |
| 19 | F/<br>35-44 | Field<br>epidemiology | 25 - 2021 | 111 | Runny nose | NS | >2180 |
| 20 | F/<br>55-64 | Laboratory | 25 - 2021 | 111 | Asymptomatic | NS | 939.1 |
| 21 | F/<br>25-34 | Field<br>epidemiology | 25 - 2021 | 111 | Asymptomatic | NS | 1912.3 |
| 22 | M/<br>25-34 | Field<br>epidemiology | 25 - 2021 | 110 | Fever, cough, sore<br>throat, adynamia,<br>headache, anosmia,<br>ageusia, arthralgia,<br>runny nose, chest pain,<br>conjunctivitis | Mu<br>(B.1.621) | >2180 |
| 23 | F/<br>25-34 | Laboratory | 26 - 2021 | 79 | Diarrhea, adynamia,<br>cough | NS | >2180 |
| 24 | F/<br>35-44 | Field<br>epidemiology | 25 - 2021 | 106 | Headache | NS | >2180 |
| 25 | M/<br>45-54 | Administrative | 28 - 2021 | 61 | Asymptomatic | Mu<br>(B.1.621) | 93.3 |
| 26 | F/<br>25-34 | Laboratory | 28 - 2021 | 62 | Fever, sore throat,<br>respiratory distress,<br>adynamia | NS | >2180 |
| 27 | F/<br>35-44 | Field<br>epidemiology | 28 - 2021 | 132 | Anosmia, ageusia, nasal<br>congestión | NS | 303.5 |
| 28 | F/<br>25-34 | Field<br>epidemiology | 25 - 2021 | 109 | Anosmia | NS | >2180 |
| 29 | F/<br>35-44 | Field<br>epidemiology | 45 - 2021 | 249 | Asymptomatic | NS | 88.1 |
| 30 | M/<br>35-44 | Field<br>epidemiology | 45 - 2021 | 249 | Asymptomatic | NS | 258.3 |
| 31 | M/<br>25-34 | Field<br>epidemiology | 48 - 2021 | 269 | Cough, sore throat,<br>adynamia | Delta<br>(AY.122) | 196.9 |

§ Anti-S IgG reported corresponds to the sample collected prior to date of the positive RT-PCR. Results are expressed as binding antibody units (BAU)/mL. Values $\geq 21.8$ BAU/ml are positive.
NS: Failed to sequencing (sequences with coverage less than 70% for lineage assignment by the Pangolin algorithm).

A total of 81.6% of the cases (31/38) had a positive RT-PCR for SARS-CoV-2. Of these, 9.7% (3/31) were detected on the same day as the first dose of the vaccine, 16.1% (5/31) between the first and second doses, and 74.2% (23/31) more than 1 week after the second dose. The remaining seven confirmed cases (18.4%) were positive for anti-nucleoprotein IgG in the serological test. Most of them (6/7) were positive after the second dose of the vaccine.

Most cases (81.6%) had IgG anti-S antibodies with values between 56 and 1600 BAU/mL when the infection occurred. The other cases (18.4%) were negative for IgG anti-S antibodies in the sample collected prior to infection confirmation; however, most of these samples were collected between days 0–21 after the first dose of vaccine (Table 2).

**Table 2.** Information for cases of SARS-CoV-2 infection confirmed by detection of IgG anti-Nucleoprotein.

| Case | Sex / Age | Occupation / activity | Date of positive result (Epi week - year) | Days after 1st dose* | Symptoms | Anti-S IgG (BAU/ml)<sup>§</sup> |
| --- | --- | --- | --- | --- | --- | --- |
| 32 | F/<br>35-44 | Laboratory | 40 - 2021 | > 90 | Asymptomatic | >2180 |
| 33 | F/<br>55-64 | Laboratory | 17 - 2021 | 0-21 | Asymptomatic | No Reactive |
| 34 | F/<br>35-44 | Administrative | 28 - 2021 | 21-60 | Asymptomatic | 1317.8 |
| 35 | M/<br>55-64 | Administrative | 32 - 2021 | 60-90 | Asymptomatic | 1694.1 |
| 36 | M/<br>45-54 | General services | 35 - 2021 | 60-90 | Asymptomatic | >2180 |
| 37 | F/<br>35-44 | Administrative | 48 - 2021 | > 90 | Headache, runny nose | >2180 |
| 38 | F/<br>55-64 | Administrative | 47 - 2021 | > 90 | Asymptomatic | >2180 |

\* Days after 1st dose are approximated according to collection date of the positive sample for IgG anti-N
§ Anti-S IgG reported corresponds to the sample collected prior to date of the positive RT-PCR. Results are expressed as binding antibody units (BAU)/mL. Values $\geq 21.8$ BAU/ml are positive.

Genomic characterization was achieved in 38.7% (12/31) of the RT-PCR-positive samples, of which 83.4% (10/12) corresponded to the variant Mu, 8.3% (1/12) Gamma, and 8.3% (1/12) Delta (Table 1).

## Discussion

Within a group of 490 healthcare workers vaccinated against COVID-19 with BNT162b2, 38 cases of SARS-CoV-2 infection were identified, of which 76.3% (29/38) occurred 7 days after the second dose and the time when full efficacy against the disease was reached [2] [10-11]. These results are consistent with previous reports on the high effectiveness of the BNT162b2 vaccine in preventing severe disease and deaths [10 - 12] but lower effectiveness against infection with the virus [4,5, 11, 13, 14].

Similar to other reports [13-14], more than 50% of the cases detected were asymptomatic, with no severe cases in infected volunteers.

Cases 4 and 19 correspond to people with a previous infection, detected by RT-PCR 6 and 14 months before detection of re-infection during this follow-up.

The Colombian third pandemic wave lasted from weeks 10 to 35 (2021), and June had the highest number of cases nationwide in 2021; hence, most cases described concur with this third pandemic peak. During this third peak, genomic surveillance showed that the most frequent variant was Mu (52.7%), followed by the Gamma and lineages (P.1, P.1.1, and P.1.2 with 23.3%) [3]. The viruses sequenced from the cases described herein concur with the predominance of the Mu variant during the third peak (Figure 1), being the variant identified in 83.4% of the samples sequenced. Additionally, the last case detected in December 2021 corresponds to the Delta variant, which started circulating in Colombia in August 2021 [15].

Finally, detection of infection was also possible through anti-N IgG serological tests in seven cases of post-vaccination infection. When the target of the vaccine is the S protein of SARS-CoV-2, the detection of antibodies against a different target, such as nucleoprotein (N), can be used to help differentiate natural infection from vaccination [16-17]. Of these seven people, only one reported having mild respiratory symptoms, while the other six never realized that they had been infected. This dataset was obtained from a well-characterized group of healthcare workers participating in a prospective cohort study that included scheduled follow-ups for collection of respiratory and blood samples and the self-report of clinical symptoms or close contact with confirmed cases of COVID-19. This allows for a very good approach to the assessment of asymptomatic infections. All findings agree with other reports in different studies that show the benefit of COVID-19 vaccines, especially against severe diseases but not against infection or re-infection.

## Data Availability

All data produced in the present study are available upon reasonable request to the authors

## Acknowledgement

The authors thank Dr. Jaime Castellanos for his review and comments, and Hector Ruiz for the figure desing.

## Funding

This research was funded by Instituto Nacional de Salud-Colombia, project code CEMIN-04-2021.

## Conflict of Interest

The authors declare no conflict of interest.

## Ethical Approval statement

Informed consent was obtained from all subjects involved in the study.

## Notes

### Competing Interest Statement

The authors have declared no competing interest.

### Funding Statement

This research was funded by Instituto Nacional de Salud-Colombia. No external funding was received.

### Author Declarations

Our "Comite de Etica y Metodologias de Investigacion" (Commitee for ethics and metodology of research) of the Instituto Nacional de Salud - Colombia gave ethical approval for this work, project code: CEMIN 04-2021

